# Double Burden of Malnutrition among Hospitalized Adults and Length of Hospital Stay in Hanoi, Vietnam: A Multicentre Prospective Cohort Study

**DOI:** 10.64898/2026.07.01.26356915

**Authors:** Tran Quyen An, Miyoshi Fumiya, Toyama Kenji, Gomi Ikuko, Nakahara Shinji, Shono Reimi, Nguyen Thuy Linh, Nguyen Thi Huong Lan, Le Thi Huong, Nakamura Teiji

## Abstract

**Background:** Evidence regarding the double burden of malnutrition (DBM) among hospitalized patients in Vietnam remains limited. This study examined nutritional status at admission and its association with length of hospital stay among adults in Hanoi.

**Methods:** This prospective observational cohort study was conducted in eight public hospitals in Hanoi between September 2018 and November 2019. Adults aged 18-60 years were assessed within 48 hours of admission using interviews, physical examination, anthropometric measurements, and medical records. Nutritional status was classified using the Subjective Global Assessment (SGA) and body mass index (BMI): undernourished (SGA-B or SGA-C or BMI <18.5 kg/m²) and overnourished (SGA-A and BMI ≥25.0 kg/m²). Length of stay was compared across nutritional-status groups using the Kruskal–Wallis test.

**Results:** Among 1,183 registered patients, 1,115 had sufficient data for analysis. Overall, 24% were undernourished and 16% overnourished. Weight loss during the preceding six months was reported by 54%, although most losses were <5%. SGA-B or SGA-C was identified in 20%, whereas 11% had BMI <18.5 kg/m². The median hospital stay was 8 days, with no significant difference across nutritional status groups.

**Conclusions:** DBM was prevalent among hospitalized adults in Hanoi. Indicators of recent nutritional deterioration were more common than low BMI, suggesting that BMI alone may overlook early disease-related nutritional decline. Nutritional status was not associated with length of stay in this relatively young, predominantly mild-to-moderate patient population. Hospital nutritional screening should therefore assess recent nutritional changes across the full BMI spectrum.

## Introduction

In recent years, the double burden of malnutrition (DBM) has emerged as a major global public health challenge, making the elimination of all forms of malnutrition a worldwide priority. The World Health Organization (WHO) defines DBM as the coexistence of undernutrition with overweight, obesity, or diet-related noncommunicable diseases within individuals, households, populations, and across the life course. Globally, this dual burden is evident in population-level data: in 2014, more than 1.9 billion adults aged 18 years and older were overweight, over 600 million were obese, with a continuously increasing trend; while 462 million were underweight.(1, 2) Similarly, among children under five years of age, an estimated 42 million were overweight, whereas 156 million experienced stunting, further highlighting the widespread and overlapping nature of malnutrition worldwide.

The DBM is also present and rapidly increasing in the Socialist Republic of Viet Nam (hereafter, Viet Nam). Among adults, the prevalence of overweight and obesity rose from 2.0% in 1992 to 5.7% in 2002, 15.6% in 2015, and 19.5% in 2021, showing a continuing upward trend.(3-6) In contrast, the prevalence of undernutrition among adults declined from 32.6% in 1992 to 24.8% in 2002, 11.6% in 2015, and 9.4% in 2021.(4, 5, 7) This coexistence of rising overnutrition and yet still-prevalent undernutrition reflects a nationwide DBM pattern, although substantial regional discrepancies persist: undernutrition remains more common in rural areas, while overweight and obesity predominate in urban areas.(4, 5, 8)

Children in Viet Nam face equally significant DBM. As of 2020, stunting affected approximately 10-15% of children under five years of age, with substantially higher prevalence in mountainous regions (25-35%).(9) In contrast, overweight and obesity were uncommon among children under five (3-5%), but affected 23% of those aged 5-9 years, rising up to 31% in urban areas. Furthermore, a longitudinal study conducted between 2009 and 2011 showed increasing trends in childhood overweight in both urban (36.9% to 38.1%) and rural areas (17.4% to 29.7%).(10)

Evidence also indicates the presence of an individual-level DBM in Viet Nam. A nationwide survey reported that micronutrient deficiencies were observed not only in underweight adult women but also in those who were overweight or obese.(11) These findings suggest that within-person DBM, in addition to between-person DBM, is also prevalent among the Vietnamese population.

Despite accumulating evidence of DBM in Viet Nam, important gaps remain in the clinical understanding of its prevalence and consequences among hospitalized patients. Most previous studies have focused on community-based epidemiological perspectives, and studies assessing nutritional status among hospitalized patients have largely been limited to those with gastrointestinal diseases.(12-14) Consequently, the overall nutritional status of hospitalized patients, the extent of DBM within healthcare facilities, and its impact on clinical outcomes remain unclear.

This study aimed to examine the nutritional status of hospitalized patients at admission in healthcare facilities in Viet Nam within the context of the DBM, to describe the prevalence of DBM, and to investigate its associations with length of hospital stay.

## Methods

### Study design

This prospective observational cohort study was conducted in eight public hospitals in Hanoi, Vietnam, from September 2018 to November 2019 to examine associations between nutritional status at admission and clinical outcomes among hospitalized patients. The present study received ethical approval from the Research Ethics Review Committee of Kanagawa University of Human Services (Decision Notice No. 71-7) and the Research Ethics Committee of Hanoi Medical University Hospital (Decision No. 35). All study participants provided written informed consent. Part of this study has been reported elsewhere.(15, 16)

### Study settings

This study was conducted across eight public hospitals in Hanoi, comprising three central-level, one provincial-level, and four district-level facilities, selected by research accessibility. All were general hospitals managing patients with diverse disease diagnoses and severities. Central and provincial hospitals provide tertiary care, while district hospitals provide secondary care. In Vietnam, public hospitals serve the majority of the population under the national health insurance scheme, whereas private hospitals operate outside this system and predominantly serve a wealthy minority.

### Participants

Eligible participants were adults aged 18–60 years who had been admitted within the preceding 48 hours, had an expected hospital stay of 3 days to 4 weeks to avoid anticipated difficulties in obtaining outcome data for those with shorter or longer stays, and who were able to stand and speak to undergo physical measurements and a questionnaire interview. Patients were excluded if they refused to participate, were deemed ineligible by their attending physician, were unable to speak Vietnamese, had infectious diseases, were bedridden, were admitted to the intensive care unit, or had not been assigned a hospital bed. In Vietnamese public hospitals, inpatient demand exceeds bed capacity, and some patients are not assigned a bed even after admission procedures; therefore, this study defined bed assignment as “hospitalized.” Of 1,183 patients enrolled across the eight hospitals, 1,115 were included in the analysis after excluding 68 patients with missing data on parameters required to determine nutritional status at admission and clinical outcomes.

### Data collection

Research assistants collected data prospectively using a standardized data form. The first assessment was conducted within 2 days of hospital admission through a structured interview, physical examinations, and data extraction from medical records. The second assessment was conducted after discharge, based on a medical record review.

The interview collected the following information: usual and current body weight (self-reported), weight changes in the preceding six months and two weeks (based on measured weight or, if unavailable, clothing fit over the preceding three months); dietary intake before admission (no change/increase, suboptimal solid diet, liquid diet, or inability to eat orally); current dietary intake (whether consuming hospital meals or outside food, and the amount consumed); gastrointestinal symptoms persisting for more than two weeks before admission (nausea, vomiting, diarrhoea, and anorexia); physical functional capacity (no dysfunction, sub-optimal function, or bedridden); and past medical history (none, malignancy, and non-malignant diseases).

Physical examination included assessment of subcutaneous fat, muscle mass, and ankle oedema, and measurement of weight and height. Weight and height were each measured twice using a portable height scale (Seca 213; Seca GmBH & Co., Hamburg, Germany) and a weight scale (BC-758; Tanita Corporation, Tokyo, Japan), and the mean of two measurements was used in the analyses.

Data extracted from the medical records at admission included the primary diagnosis (cause of admission). Post-discharge information from the medical record included the discharge date, disposition (discharged to home or transferred to another facility), and clinical outcome (recovery, remission, unchanged, worsened, anticipated death, improved, or unknown).

The research assistants were from the nutrition department staff of each hospital. To standardize the interview and physical measurements across sites, trained Vietnamese dieticians provided them with training in interview procedures and physical measurement techniques before starting data collection.

### Variables

Age was categorized as 18–24, 25–34, 35–44, 45–54, and ≥ 55 years. The primary diagnosis was coded and categorized according to the International Statistical Classification of Diseases and Related Health Problems, Tenth Revision (ICD-10), using chapter-level categorization. Diagnoses with small case counts were combined into an "other" category.

### Nutrition assessment

After obtaining the data, the research assistants assessed participants’ nutritional status at hospital admission. Percentage weight loss was calculated based on the self-reported usual weight and measured body weight as follows:

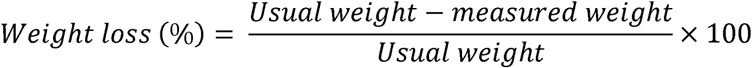

Metabolic demand was assessed based on the primary diagnosis. Conditions with high metabolic demand included severe trauma, severe inflammatory diseases, severe infections, and those requiring major surgery.

Overall nutritional status was assessed using the Subjective Global Assessment (SGA), developed by Baker et al.,(17) to assess malnutrition risk. It incorporates weight change, change in dietary intake, gastrointestinal symptoms, physical functions, and metabolic stress. Its validity has been assessed against other screening tools and objective indicators.(17-19) Whereas SGA ratings are determined by a subjective weighing of all collected information rather than an explicit numerical scoring system, their reliability across multiple evaluators has also been demonstrated.(17, 20) The SGA classifies the risk of malnutrition into three categories, as follows:

SGA-A (normal nutrition): less than 5% weight loss, or 5–10% loss with recent stabilization or regain in two or more weeks before the admission (if not weighed, not feeling loose clothes); no or minimal loss of fat/muscle; and normal to minimally impaired functional capacity.
SGA-B (mild to moderate malnutrition): ongoing weight loss of 5–10% (if not weighed, slight looser clothing was used as a proxy indicator); mild to moderate compromised food intake; gastrointestinal symptoms; mild to moderate loss of fat/muscle; slight oedema; and mild to moderate functional impairment.
SGA-C (severe malnutrition): ongoing weight loss of over 10% (if not weighed, much looser clothing was used as a proxy indicator), weight loss in the past two weeks, obviously compromised food intake, severe loss of fat/muscle, marked oedema, and moderate to severe functional capacity.

Body mass index (BMI) at admission was calculated from measured weight and height as follows:

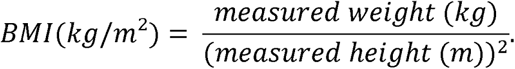

BMI was categorized as <18.5 kg/m² (undernutrition), 18.5 to <25.0 kg/m² (normal nutrition), and ≥25 kg/m² (overnutrition). The lower threshold aligns with international standards, while the upper threshold adheres to the 25.0 kg/m² cutoff for obesity as recommended for Asian populations.(21, 22)

Nutritional status at admission was classified into three categories based on the SGA and BMI: undernourished (SGA-B or SGA-C, or BMI < 18.5 kg/m²); well-nourished (SGA-A and BMI 18.5 to <25.0 kg/m²); overnourished (SGA-A and BMI ≥ 25 kg/m²).

### Outcome variable

In the analysis to investigate the association between the nutritional status and prognosis, we used length of hospital stay as the primary outcome. Unfavourable outcomes at discharge (worsened conditions or anticipated death) occurred too infrequently to allow separate analysis.

### Statistical analysis

Patient characteristics, including the prevalence of overnutrition and undernutrition, were summarized as proportions and medians with interquartile ranges. The Kruskal-Wallis test was employed to assess the association between the three nutrition categories (overnutrition, undernutrition, and normal nutrition) and length of hospital stay, with a significance level of 0.05.

Sample size was determined based on a pilot study conducted in January 2018, which estimated the prevalence of malnutrition (including both over- and undernutrition) at 25%. The required sample size was 1,153 to constrain the 95% confidence interval within ±2.5%. With this sample size, a power of 0.8, and a significance level of 0.05, the minimum detectable effect size (Cohen’s W) in a 3×2 contingency table with 2 degrees of freedom was 0.095, indicating a small effect size.

## Results

Of the 1,183 enrolled patients, middle-aged adults comprised the majority, with a nearly balanced sex distribution (Table 1). Most patients had no functional impairment. Non-malignant comorbidities were prevalent, such as gastrointestinal disorders, diabetes, and cardiovascular disease. The most common primary diagnoses at discharge were gastrointestinal diseases (21%), followed by neoplasms. Overall, 28% of patients reported gastrointestinal symptoms, such as anorexia and nausea.

**Table 1.** Patient characteristics (n = 1,183)

| **Characteristic** | **n (%)** |
| --- | --- |
| **Age** |  |
| < 25 years | 94 (8) |
| 25-34 years | 167 (14) |
| 35-44 years | 263 (22) |
| 45-54 years | 325 (27) |
| 55-60 years | 334 (28) |
| **Sex** |  |
| Female | 583 (52) |
| Male | 531 (48) |
| Data missing | 69 |
| **Functional Capacity** |  |
| No dysfunction | 920 (82) |
| Working sub-optimally/ ambulatory | 206 (18) |
| Bed-ridden | 1 (<0.5) |
| Data missing | 56 |
| **Medical History <sup>a</sup>** |  |
| Nothing/unknown | 362 (31) |
| Malignancy | 44 (4) |
| Non-malignancy | 769 (65) |
| Non-malignancy breakdown |  |
| Diabetes | 173 (15) |
| Renal | 71 (6) |
| Cardiovascular | 166 (14) |
| Cerebrovascular | 37 (3) |
| Pulmonary | 45 (4) |
| Hepatic | 113 (10) |
| Gastrointestinal | 204 (17) |
| Other | 267 (23) |
| Data missing | 8 |
| **Primary diagnosis at discharge** |  |
| Gastrointestinal diseases | 247 (21) |
| Neoplasms | 199 (17) |
| Genitourinary diseases | 119 (10) |
| Endocrine, nutritional, and metabolic diseases | 118 (10) |
| Otolaryngological diseases | 75 (6) |
| Cardiovascular diseases | 68 (6) |
| Infectious diseases | 60 (5) |
| Respiratory diseases | 46 (4) |
| Musculoskeletal or connective tissue diseases | 33 (3) |
| Injuries (consequences of external causes) | 29 (2) |
| Hematological or immunological diseases | 23 (2) |
| Neurological diseases | 22 (2) |
| Ophthalmological diseases | 14 (1) |
| Dermatological disease | 9 (1) |
| Other diseases<sup>b</sup> | 108 (9) |
| Data missing | 13 |
| **Gastrointestinal Symptoms** |  |
| No symptom | 851 (72) |
| Symptoms existed | 332 (28) |
| Breakdown of symptoms <sup>c</sup> |  |
| Nausea | 183 (15) |
| Vomiting | 82 (7) |
| Diarrhea | 55 (5) |
| Loss of Appetite (Anorexia) | 201 (17) |

<sup>a</sup> Multiple diseases may be selected
<sup>b</sup> This category includes neonatal, perinatal, and psychiatric diseases, as well as abnormal findings not elsewhere classified
<sup>c</sup> Some patients had multiple symptoms

Table 2 presents the nutritional status of the participants. The majority (54%) reported weight loss during the preceding six months, of whom 71% experienced a loss of less than 5%. Over the preceding two weeks, 79% reported no or unknown weight change. In addition, one quarter reported that their clothes had become looser over the preceding three months.

**Table 2.** Nutritional status and dietary intake (n = 1,183)

| **Characteristic** | **n (%)** |
| --- | --- |
| **Weight Change in the past 6 Months** |  |
| No change/ Increase/Unknown | 535 (46) |
| Decrease | 640 (54) |
| Data Missing | 8 |
| **Weight Loss in 6 months (n = 640), kg, median (IQR)** | 2.1 (1.0 – 5.3) |
| **Weight loss proportion (n = 640)** |  |
| 0–< 5% loss | 453 (71) |
| 5–10% loss | 119 (19) |
| > 10% loss | 68 (11) |
| **Weight Change in the past 2 Weeks** |  |
| No change/ Unknown | 920 (79) |
| Increase | 38 (3) |
| Decrease | 211 (18) |
| Data Missing | 14 |
| **Weight loss in 2 weeks (n = 124 <sup>a</sup>), kg, median (IQR)** | 2.0 (1.0 – 2.0) |
| **Felt clothes becoming looser in the past 3 months** |  |
| No change/ Do not notice | 879 (74) |
| Become bit looser | 233 (20) |
| Become much looser | 69 (6) |
| Data Missing | 2 |
| **Changes in Dietary Habits** |  |
| No change/ increase | 928 (81) |
| Suboptimal solid diet | 161 (14) |
| Liquid diet | 37 (3) |
| Starvation | 14 (1) |
| Data Missing | 43 |
| **Duration of Dietary Change (n = 159<sup>b</sup>), weeks median (IQR)** | 2 (1 – 4) |
| **Current Diet Type** |  |
| Hospital meal | 221 (19) |
| Meal from outside | 937 (81) |
| Data Missing | 25 |
| **Hospital Meal Intake (n=176)<sup>c</sup>** |  |
| < 50% | 11 (6) |
| 50% – 75% | 42 (24) |
| > 75% | 123 (70) |
| **High Metabolic Demand Condition** |  |
| No | 1,009 (94) |
| Yes | 66 (6) |
| Data Missing | 108 |
| **Subcutaneous Fat Reduction** |  |
| Normal | 992 (84) |
| Mild/moderate | 180 (15) |
| Severe | 7 (1) |
| Data Missing | 4 |
| **Muscle Wasting** |  |
| Normal | 1,028 (87) |
| Mild/moderate | 147 (12) |
| Severe | 4 (<0.5) |
| Data Missing | 4 |
| **Ankle Edema** |  |
| Normal | 1,147 (98) |
| Mild/moderate | 26 (2) |
| Severe | 2 (<0.5) |
| Data Missing | 8 |
| **Subjective Global Assessment Score** |  |
| A | 946 (80) |
| B | 200 (17) |
| C | 30 (3) |
| Data Missing | 7 |
| **BMI** |  |
| < 18.5kg/m<sup>2</sup> | 131 (11) |
| 18.5 to < 25kg/m<sup>2</sup> | 862 (73) |
| ≥ 25kg/m<sup>2</sup> | 190 (16) |

<sup>a</sup> Among the 211 patients who reported weight decrease in the past 2 weeks, 87 did not report the decreased amount
<sup>b</sup> Among the 212 patients who reported dietary changes, 53 did not report the duration
<sup>c</sup> Among the 221 patients who were taking hospital meals, 45 did not answer the amount they took

Dietary intake was largely maintained, with 19% reporting a decline. Among participants who reported dietary change and provided its duration, the median duration was two weeks. Only one-fifth of participants consumed hospital meals; of those with available intake data, 70% consumed more than 75% of their meals.

The clinical nutrition assessment indicated that most patients had normal metabolic and nutritional status. A small proportion exhibited high metabolic demand, subcutaneous fat loss, and muscle wasting. Based on SGA, 20% were at high risk of malnutrition (SGA-B or C). Based on BMI, 11% were underweight (BMI<18.5 kg/m²) and 16% were overweight (BMI≥25.0 kg/m²).

Overall, patient clinical outcomes were favourable (Table 3). Among 1122 patients with available data, the median hospital stay was 8 days, with the majority being discharged within two weeks. Most patients were discharged after recovery.

**Table 3.**
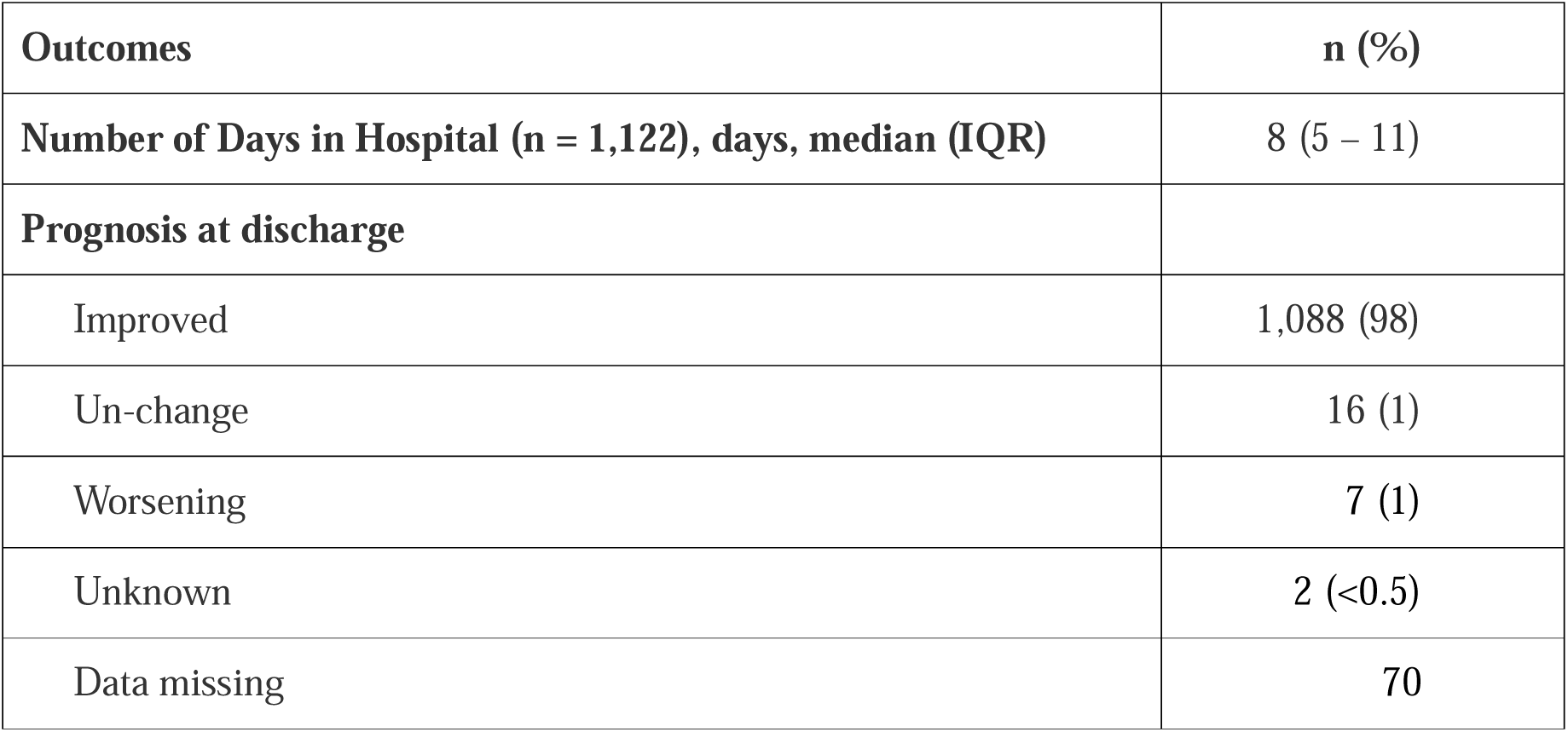
Clinical Outcomes (n = 1,183)

| **Outcomes** | **n (%)** |
| --- | --- |
| **Number of Days in Hospital (n = 1,122), days, median (IQR)** | 8 (5 – 11) |
| **Prognosis at discharge** |  |
| Improved | 1,088 (98) |
| Un-change | 16 (1) |
| Worsening | 7 (1) |
| Unknown | 2 (<0.5) |
| Data missing | 70 |

Table 4 presents the association between nutritional status and length of hospital stay. Among 1,115 patients with available data, 24% were classified as undernourished and 16% as overnourished based on SGA and BMI. Length of stay was similar across nutritional status groups.

**Table 4.** Association between nutritional status category and length of hospital stay (n = 1,115)

| Nutritional status <sup>a</sup> | n (%) <sup>b</sup> | Length of hospital stay (days) | P-value <sup>c</sup> |
| --- | --- | --- | --- |
| Undernutrition | 267 (24) | 8 (5-12) | 0.229 |
| Normal nutrition | 674 (60) | 8 (5-10) |  |
| Overnutrition | 174 (16) | 8 (5-10) |  |

<sup>a</sup> Determined based on SGA and BMI
<sup>b</sup> Data missing in 68 patients
<sup>c</sup> Kruskal-Wallis test

## Discussion

This study identified two important nutritional problems among hospitalized adults in Hanoi. First, undernutrition (24%) and overnutrition (16%) coexisted at the hospital level according to the combined SGA and BMI criteria. Second, recent deterioration in nutrition was common, as indicated by weight loss over the preceding six months and physical signs of nutritional depletion, including subcutaneous fat loss and muscle wasting, which BMI alone could not capture. However, nutritional status was not significantly associated with length of hospital stay.

This DBM pattern is consistent with a health system undergoing a nutritional and epidemiological transition,(23, 24) in which hospital nutrition services must provide appropriate care for both ends of the nutritional spectrum. Undernourished patients require early nutritional support to prevent further tissue loss and functional deterioration, whereas those with overweight or obesity require nutrition care that addresses long-term cardiometabolic risk, with close attention to recent reductions in food intake, weight, or muscle mass. Thus, hospital nutrition services should be designed to accommodate the DBM.

The second finding highlights the importance of dynamic nutritional assessment rather than relying solely on static measures, such as BMI, to identify the risk of malnutrition among hospitalized patients. Although most patients were within the normal BMI range, many reported recent weight loss or showed signs of tissue depletion, suggesting that BMI alone may fail to identify patients with early or disease-related malnutrition. Particularly in acute care settings, acute illness, poor dietary intake, gastrointestinal symptoms, and treatment-related stress may rapidly deteriorate nutritional status and lead to recent weight loss even before noticeable changes in BMI become apparent.(25-27) This issue is particularly relevant in the context of increasing overweight and obesity,(1, 6) as a considerable amount of weight loss would be required for their BMI to fall to the underweight category. Therefore, nutritional assessments should incorporate weight trajectory, dietary intake, clinical symptoms, functional status, and physical signs of tissue depletion to complement BMI.(28, 29)

In contrast to some previous studies, nutritional status was not significantly associated with length of hospital stay in this cohort.(30, 31) The most plausible explanation is the restricted clinical and nutritional severity of the study population. Patients who were bedridden, admitted to an intensive care unit, unable to stand or participate in an interview, or affected by certain severe conditions were excluded because the study required interviews and anthropometric measurements. Consequently, most participants had mild-to-moderate disease conditions, relatively preserved functional capacity, and predominantly mild nutritional deterioration. Clinical outcomes were also generally favourable, and the median hospital stay was relatively short, limiting the variation in both nutritional status and clinical outcomes, which may have reduced the study’s ability to detect a statistically significant association.

Length of stay may also be affected by non-clinical factors that were not measured, including hospital discharge practices, availability of post-discharge care, administrative procedures, and preferences of patients and their families. For example, some families may prefer extended hospitalization until they perceive full recovery, even when it is medically unnecessary. These factors may have introduced additional variation in length of stay and obscured any association with nutritional status.

Therefore, the null finding should not be interpreted as evidence that malnutrition does not affect outcomes. The findings are likely to reflect the characteristics of this study population, whereas the association may appear among older, more severely ill, bedridden, or critically ill patients who were not represented in this study.

The high prevalence of overnutrition as well as undernutrition revealed in this study supports the need for routine nutritional screening at hospital admission for patients across the full BMI spectrum. Screening should include dynamic indicators such as recent weight loss, changes in dietary intake, gastrointestinal symptoms, functional capacity, and physical signs of fat or muscle depletion, rather than relying solely on BMI. Patients at risk of undernutrition, regardless of their current BMI, should receive timely dietitian assessment, symptom management, appropriate nutritional support, and ongoing monitoring. At the same time, the substantial prevalence of overnutrition indicates a need for hospital nutrition services to address obesity and cardiometabolic risk. Addressing the DBM in Vietnamese hospitals will require a comprehensive approach that goes beyond conventional undernutrition-focused care.

Several limitations should be considered when interpreting these findings. First, the eligibility criteria excluded patients older than 60 years and those who were bedridden, critically ill, unable to stand, or otherwise unable to complete the interview and physical measurements. These groups are likely to have a higher prevalence and greater severity of malnutrition. Their exclusion may therefore have led to a conservative estimate, and the true burden of DBM in Vietnam may be substantially higher; it may have also attenuated the association between nutritional status and length of stay. The findings should consequently not be generalized to older or severely ill hospital populations.

Second, some information on usual weight, previous weight loss, dietary intake, and gastrointestinal symptoms was self-reported and may be subject to recall or reporting bias. Several measures were taken to minimize this problem: research assistants used a standardized interview format and received training from experienced Vietnamese dietitians; changes in clothing fit were used as a proxy for weight change when measured weight was unavailable; self-reported information was interpreted alongside objectively measured weight, height, and clinical examination of subcutaneous fat and muscle mass. These procedures reduced, although did not completely eliminate, the possibility of measurement error. Any remaining error was likely to reduce the distinction between nutritional groups and therefore weaken rather than exaggerate the observed associations.

Third, the length of hospital stay was the only outcome examined and may not adequately capture all clinically relevant effects of malnutrition, as it is also influenced by non-clinical factors such as discharge and post-discharge practices and family preferences, which were not measured in this study. Outcomes such as complications, infection, functional decline, readmission, mortality, and post-discharge recovery may be more sensitive to nutritional status and should be addressed in future studies.

Finally, the comparison of length of stay was unadjusted for hospital-level and individual-level factors. Future studies should include a broader spectrum of patients and use adjusted analyses to better examine the association between nutritional status and patient outcomes.

## Conclusions

This multicentre study demonstrates a high prevalence of both undernutrition and overnutrition among hospitalized patients in Hanoi, Vietnam; recent weight loss, reduced dietary intake, gastrointestinal symptoms, and physical signs of tissue depletion were more common than low BMI. Nutritional status was not associated with length of hospital stay, reflecting the relatively young and predominantly mild-to-moderate severity of the study population. These findings highlight the need for comprehensive nutritional screening across the full BMI spectrum, incorporating dynamic indicators rather than BMI alone. Future studies should examine more diverse patient populations and clinically sensitive outcomes.

## Data Availability

All data produced in the present study are available upon reasonable request to the authors

## Presentation history

Part of this research was presented at the 8th Asian Congress of Dietetics held from 19 to 21 August 2022 in Yokohama, Japan.

Part of this research was presented as the second author’s master’s thesis at the Graduate School of Health and Social Services at Kanagawa University of Human Services in 2021.

## Data statement

The data that support the findings of this study are available on request from the corresponding author. The data are not publicly available due to restrictions on their containing information that could compromise the privacy of research participants.

## Ethical considerations

The present study received ethical approval from the Research Ethics Review Committee of Kanagawa University of Human Services (Decision Notice No. 71-7) and the Research Ethics Committee of Hanoi Medical University Hospital (Decision No. 35). All study participants provided written informed consent.

## Declaration of AI use in the manuscript preparation

During the preparation of this manuscript, the authors used ChatGPT, Claude, and Grammarly in order to select appropriate terms, correct grammatical errors, improve readability, and design the table. After using them, the authors reviewed and edited the content as needed and take full responsibility for the content of the article.

## Author contributions

**TRAN Quyen An**: Formal analysis, Writing - original draft, and Visualization; **MIYOSHI Fumiya**: Formal analysis, Investigation, Data curation, and Writing - original draft; **TOYAMA Kenji**: Conceptualization, Methodology, Investigation, Writing - review and editing, Supervision, and Project administration; **GOMI Ikuko**: Conceptualization, Methodology, Investigation, Writing - review and editing, Supervision, and Project administration; **NAKAHARA Shinji**: Conceptualization, Methodology, Investigation, Writing - review and editing, Visualization, Supervision, and Project administration; **SHONO Reimi**: Data curation and Writing - review and editing; **NGUYEN Thuy Linh**: Methodology, Investigation, Writing - review and editing, and Project administration; **NGUYEN Thi Huong Lan**: Methodology, Investigation, Writing - review and editing, and Project administration; **LE Thi Huong**: Methodology, Writing - review and editing, Supervision, and Project administration; **NAKAMURA Teiji**: Methodology, Writing - review and editing, Supervision, Project administration, and Funding acquisition.

